# A geopositioned, evidence-graded, pan-species compendium of Mayaro Virus occurrence

**DOI:** 10.1101/2023.03.07.23286930

**Authors:** Michael Celone, Alex Potter, Barbara A. Han, Sean P. Beeman, Bernard Okech, Brett Forshey, James Dunford, George Rutherford, Neida Karen Mita-Mendoza, Elizabet Lilia Estallo, Ricardo Khouri, Isadora Cristina de Siqueira, Kyle Petersen, Ryan C. Maves, Assaf Anyamba, Simon Pollett

## Abstract

Mayaro Virus (MAYV) is an emerging health threat in the Americas that can cause febrile illness as well as debilitating arthralgia or arthritis. To better understand the geographic distribution of MAYV risk, we developed a georeferenced database of MAYV occurrence based on peer-reviewed literature and unpublished reports. Here we present this compendium, which includes both point and polygon locations linked to occurrence data documented from its discovery in 1954 until 2022. We describe all methods used to develop the database including data collection, georeferencing, management and quality-control. We also describe a customized grading system used to assess the quality of each study included in our review. The result is a comprehensive, evidence-graded database of confirmed MAYV occurrence in humans, non-human animals, and arthropods to-date, containing 266 geo-positioned occurrences in total. This database - which can be updated over time - may be useful for local spill-over risk assessment, epidemiological modelling to understand key transmission dynamics and drivers of MAYV spread, as well as identification of major surveillance gaps.

## Background & Summary

First detected in Trinidad in 1954 ^1^, Mayaro virus (MAYV) is a mosquito-borne *Alphavirus* that causes periodic outbreaks of febrile illness in several Latin American countries ^2-5^. Serological surveys conducted in the Americas suggest widespread circulation of the virus throughout the region ^6-8^. Although MAYV may cause debilitating arthralgia or arthritis, it often presents with non-specific symptoms that may be clinically indistinguishable from other vector-borne diseases such as dengue or Zika ^9^. No licensed vaccine or antiviral treatment currently exists, and the current standard of care for MAYV infections is limited to supportive care ^9,10^.

Collating and evaluating the current evidence regarding the distribution of MAYV occurrence is a critical step in characterizing its transmission potential and identifying the communities at greatest risk for MAYV outbreaks. Such compendium databases have proven valuable for the study and prevention of other emerging and re-emerging pathogens and diseases, such as Middle East respiratory syndrome coronavirus (MERS-CoV), leishmaniasis, Crimean-Congo hemorrhagic fever, and dengue viruses ^11-14^.

Previously published reviews have explored MAYV occurrence in the Americas ^15,16^. This review seeks to fill gaps in the current literature by providing the highest possible level of geographic resolution for the available MAYV occurrence data across humans, non-human animals, and arthropods, coupled with an evidence-based grading for each of those occurrences. In addition to drawing from occurrences identified in a human systematic review, this compendium database includes occurrences identified in another systematic review ^17^ evaluating the distribution of MAYV occurrence among non-human animal and arthropod species in the Americas. Collectively, these two systematic reviews provide a comprehensive compendium of MAYV occurrence to permit ecological and epidemiological risk prediction and forecasting. Further, these two systematic reviews put forward joint citable frameworks for the field to evaluate the quality of studies that propose non-human animal, arthropod, and human MAYV occurrence.

This georeferenced, evidence-graded MAYV database contains 266 unique localities across 15 countries published between 1954 and 2022. The methods described below are adapted from previously published disease occurrence compendiums ^11-14^.

## Methods

### Data Collection

Data collection and abstraction for non-human animal and arthropod data was described in a previously published study ^17^. We followed a similar data collection strategy for human MAYV occurrence data. We first searched Embase, Web of Science, PubMed/MEDLINE, and SciELO databases for English, Spanish, and Portuguese language articles published between January 1954 and January 2021 using the search term “Mayaro”. Furthermore, a PubMed/MEDLINE alert using the search term “Mayaro” captured five additional eligible studies reporting human MAYV occurrence that were published between the initial search and May 2022.

This database search was extended using the bioRxiv (https://www.biorxiv.org/) and medRxiv (https://www.medrxiv.org/) pre-print databases. We complemented these database search results with ‘grey literature’, including hand-searched bibliographies of the included articles and MAYV review articles (including systematic reviews), the Pan American Health Organization (PAHO) Institutional Repository for Information Sharing database (iris.paho.org), the GIDEON database (https://www.gideononline.com/), and GenBank [29] (https://www.ncbi.nlm.nih.gov/genbank/). We also searched for dissertations from several Brazilian university repositories including Instituto Evandro Chagas (https://patua.iec.gov.br/), Universidade de São Paulo (https://teses.usp.br/), Universidade Federal de Goiás (https://repositorio.bc.ufg.br/), Pontifícia Universidade Católica do Rio Grande do Sul (http://tede2.pucrs.br/), Universidade Federal de Mato Grosso (https://bdm.ufmt.br/), Fundação Oswaldo Cruz (https://portal.fiocruz.br/), and Universidade Federal do Pará (http://repositorio.ufpa.br/). In addition, we searched conference handbooks that are available online (2004-2019) from the American Society of Tropical Medicine and Hygiene (https://www.astmh.org/annual-meeting/past-meetings). Articles were considered for eligibility if they reported original research studies on MAYV occurrence in humans including serological surveys, outbreak investigations, case reports, or surveillance studies. After the initial literature search, we conducted a secondary search to identify any relevant articles describing the occurrence of Uruma virus. Initially described as a novel human pathogen in 1959 ^18^, Uruma virus is now considered a strain of MAYV ^19^. Therefore, it was decided that Uruma virus records would be included in our systematic review.

Two reviewers independently screened all titles and abstracts to determine articles that could immediately be discarded based on relevancy and articles that should be included in the second stage of review. Results were compared to reconcile any differences between the reviewers. Each reviewer then independently read the full text of potentially eligible articles identified through screening and identified articles that were candidates for inclusion in the study. Results were compared to reconcile any differences between the two reviewers. A third-party reviewer adjudicated if consensus was not reached between the reviewers. From those studies deemed eligible, data was extracted from articles by one reviewer using a predetermined data abstraction tool in Microsoft Excel. Five percent of entries were randomly selected and reviewed by a second reviewer.

Overall, 144 research items (including journal articles, dissertations, news articles, GenBank entries, etc.) were deemed eligible. All eligible articles are included in the Supplementary materials (Data Citation 1). No additional articles were deemed eligible following the secondary search for Uruma virus.

### Grading quality of evidence

For human occurrences, we developed a customized grading system to assess the quality of each study included in our review. This followed a similar framework we developed for evaluating the quality of each study included in the published systematic review on MAYV occurrence in non-human animals and arthropods ^17^. We assigned each study in our systematic review a grade for each of four quality items: clarity of research question/objective; description of study methods; description of sampling methods; and validity of diagnostic tests. For each quality item, eligible studies were assigned a score of 3 (strong evidence), 2 (moderate evidence), 1 (weak evidence), or unable to judge. Studies were deemed unable to judge if the information provided was insufficient to assign quality scores (e.g., a single GenBank entry, conference abstract, or report). Table 1 refers to a description of the four quality grading domains.

**Table 1:** Evidence quality grading scheme – Human infection.

| **Quality item** | **3-Strong** | **2- Moderate** | **1- Weak** | **Unable to judge** |
| --- | --- | --- | --- | --- |
| **Was the research question/objective clearly described and stated?** | Described in full detail | Described in moderate detail (neither strong nor weak evidence) | Not described at all | Unable to judge based on the information presented |
| **Were the study design (including location and year of study) and participant recruitment presented in a reproducible way?** | Described in full detail | Described in moderate detail (neither strong nor weak evidence) | Not described at all | Unable to judge based on the information presented |
| **Was relevant information collected on participants, including travel history, occupation, location of residence, etc.?** | Described in full detail | Described in moderate detail (neither strong nor weak evidence) | Not described at all | Unable to judge based on the information presented |
| **Was MAYV exposure positivity measured in a valid way?<sup>1</sup>** | PCR or viral culture or high-specificity serological method (e.g., PRNT) | Lower -specificity serological assay (i.e., HI and ELISA) without exclusion of sero-cross-reactivity | No confirmatory assay, presumptive exposure only | Unable to judge based on the information presented<sup>2</sup> |

MAYV: Mayaro virus; PCR: polymerase chain reaction; PRNT: plaque-reduction neutralization test; HI: hemagglutination inhibition; ELISA: enzyme-linked immunosorbent assay
<sup>1</sup>When multiple methods were used with varying validity (e.g., HI and PCR), we assigned a score to the most valid assay that detected MAYV positivity.
<sup>2</sup>When no MAYV-positive results were found, we assigned a score of N/A.

**Table 2.** Unique occurrences by country and location type.

| **Country** | **Point** | **Polygon** | **Total** |
| --- | --- | --- | --- |
| **Brazil** | 35 | 100 | 135 |
| **Peru** | 10 | 24 | 34 |
| **French Guiana** | 23 | 8 | 31 |
| **Suriname** | 5 | 9 | 14 |
| **Trinidad & Tobago** | 11 | 2 | 13 |
| **Colombia** | 3 | 6 | 9 |
| **Venezuela** | 4 | 3 | 7 |
| **Panama** | 4 | 2 | 6 |
| **Bolivia** | 1 | 5 | 6 |
| **Ecuador** | 0 | 5 | 5 |
| **Mexico** | 0 | 2 | 2 |
| **Haiti** | 0 | 1 | 1 |
| **Costa Rica** | 0 | 1 | 1 |
| Antigua | 0 | 1 | 1 |
| Guyana | 0 | 1 | 1 |
| TOTALS | 96 | 170 | 266 |

**Table 3.** Mayaro virus occurences by host type and spatial resolution.

|  | Points | Custom | Admin3 | Admin2 | Admin1 | Admin0 |
| --- | --- | --- | --- | --- | --- | --- |
| Human | 74 | 24 | 7 | 88 | 19 | 10 |
| Non-human animal | 7 | 2 | 0 | 20 | 4 | 1 |
| Arthropod | 15 | 3 | 1 | 2 | 1 | 2 |

Two reviewers independently graded the evidence quality for each study and results were compared to reconcile any differences between the two reviewers. A third-party reviewer adjudicated if the two initial reviewers did not reach consensus.

### Geo-positioning of the MAYV Occurrence Data

All available location information from each confirmed MAYV case was extracted from each article and added to the database based on methods that have been described previously ^11-14^. Each occurrence of MAYV was designated as either a point or polygon location according to the spatial resolution provided in the article. When specific latitude and longitude coordinates were provided, they were verified in Google Maps (https://www.google.com/maps) and added to the database as a point location and coordinates were converted to decimal degrees. If a neighborhood, town, village, or small city was explicitly mentioned in the article and the uncertainty of the location was less than 5 kilometers (km), it was entered into the database as a point location and its centroid coordinates were recorded. An online gazetteer (www.geonames.com) as well as Google Maps or ArcGIS (ESRI 2011. ArcGIS Pro: Release 2.6.0. Redlands, CA: Environmental Systems Research Institute), along with contextual information, were used to verify site locations.

For studies that reported a less precise spatial resolution such as states or counties, first level (ADM1), second level (ADM2) or third level (ADM3) administrative divisions were recorded as polygons and coded according to the global administrative unit layers (GAUL) from the Food and Agriculture Organization ^20^. If the uncertainty of a specific named location was greater than 5 km (e.g., the city of Manaus, Brazil), the occurrence was assigned to a custom polygon created in ArcGIS that encompassed the extent of the city. In the rare case that no specific intra-country location was provided, the record was assigned to its country of occurrence (ADM0). When place names were duplicated (i.e., the ADM1 and ADM2 units had the same name), the coarsest spatial resolution was used. For example, if the MAYV case location was reported as “*Cusco, Peru,”* with no additional information provided, the record was assigned to the Cusco ADM1 polygon. However, if the study specified that the case occurred in the “*City of Cusco”*, the record was assigned to a custom polygon that encompassed the City of Cusco. The centroid coordinates of ADM1, ADM2, and ADM3 polygons, or custom polygons were retrieved from the GeoNames gazetteer whenever possible. If centroid coordinates were not available in GeoNames, they were estimated using Google Maps. The coordinates for each georeference and the methods and source used to obtain the coordinates were documented in the compendium.

Uncertainty was measured in kilometers for each MAYV occurrence point or polygon ^21^. For polygons, uncertainty was calculated as the distance from the polygon centroid coordinates to the polygon’s furthest boundary. For point locations with well-defined boundaries, the same procedure was followed, whereby the uncertainty encompassed the extent of the location’s area. When locations did not have well defined boundaries, uncertainty was calculated as half the distance to the nearest named place ^21,22^. Calculation of uncertainty was completed using measurements from Google Maps. When authors provided exact coordinates gathered using GPS, uncertainty was calculated using a georeferencing calculator ^23^. Exact coordinates were only used if authors provided a high level of precision (e.g., precision higher than “minutes” in degrees-minutes-seconds format and similarly high precision for coordinates in decimal degrees). When coordinates were provided at a low precision, we georeferenced the named place instead.

Several articles reported the diagnosis of MAYV in human blood samples at urban hospitals. If no relevant information was provided on the study participants (e.g., place of residence), we georeferenced the ADM2 unit in which the hospital was located.

### Data and Metadata Records

This database is available in the Supplementary materials (Data Citation 1). Each of the 280 rows represents a unique occurrence of MAYV in a human, non-human animal, or arthropod. Location IDs for points and polygons were assigned to each unique location. The MAYV occurrence database contains the following fields, following best-practice nomenclature as previously documented in georeferenced compendiums of other pathogens ^11-14^:

1. **Location_ID:** A unique identifier was assigned to each georeference. The prefix used in the location ID denoted the georeference type: *ADM 0, 1, 2*, or *3* for administrative units, *CP* for custom polygons, and *P* for point locations. Separate studies with duplicate georeferences were assigned the same Location ID, and duplicates were removed according to the methods described above.
2. **Author_Year:** The first author and publication year for each record.
3. **Ref_Number:** A reference identification number was documented when applicable. A PubMed ID number was recorded for all published studies. If this was not available a DOI, GenBank locus, URL, ProMED identifier, etc., was captured.
4. **Year_MAYV_Start:** The earliest year that MAYV infection was detected within the publication was recorded if available. If studies only included a range of years and did not specify the precise year that MAYV was found, this range was documented. Note that this variable refers to infection detection and doesn’t infer the onset of infection (particularly in the case of serological-based occurrence studies).
5. **Year_MAYV_End:** The latest year that MAYV infection was detected within the publication was recorded if available. If studies only included a range of years and did not specify the precise year that MAYV was found, this range was documented. We followed the methods of Hill et al.,^24^ when studies did not report any year (i.e., an assumption was made that the case was detected three years before publication).
6. **Host_Type:** One of three host types was documented for each occurrence: human, non-human animal, or arthropod. If multiple host types were detected with MAYV in the same location, a separate row was included for each host type.
7. **Location_Description:** We documented relevant information related to the location of the occurrence record. This field included the decisions made during the georeferencing process to reach the final determination regarding the location of each record.
8. **Adm0:** The country where MAYV occurrence was detected.
9. **Adm1:** The first level administrative unit where MAYV occurrence was detected (if available).
10. **Adm2:** The second level administrative unit where MAYV occurrence was detected (if available).
11. **GAUL_code:** When a MAYV occurrence was georeferenced as an ADM1 or ADM2 administrative polygon, the GAUL code was included.
12. **Finer_Res:** If finer spatial resolution was documented (e.g., a town, city, or exact coordinates) this was recorded.
13. **Location_Type**: Each occurrence was documented as either a point or polygon location type, depending on the spatial resolution that was provided. Custom polygons are available as shapefiles in the Supplementary materials (Data Citation 1). These can be opened in GIS software or using statistical packages that handle spatial data.
14. **Admin_Level:** The administrative level for each polygon location was recorded as either 0 (country level), 1 (first level administrative division), 2 (second level administrative division), or 3 (third level administrative division) depending on the spatial resolution that is provided. If the occurrence was georeferenced as a point location or custom polygon, -999 was recorded.
15. **Y_Coord:** The longitude coordinate was recorded in decimal degrees. The coordinates were taken verbatim from the article when available. Otherwise, the polygon centroid was recorded.
16. **X_Coord:** The latitude coordinate was recorded in decimal degrees. The coordinates were taken verbatim from the article when available. Otherwise, the polygon centroid was recorded.
17. **Coord_Source:** This field describes how the coordinates were determined. Possibilities include the following:
  a. Exact coordinates provided in the article.
  b. Polygon centroid coordinates retrieved from GeoNames.
  c. Location was determined based on the details provided in the article (e.g., a specific neighborhood was mentioned), and centroid coordinates were subsequently determined using Google Maps.
18. **Uncertainty_km**: The amount of uncertainty associated with the record, measured in km.
19. **Uncertainty_Description:** The method used to calculate uncertainty for each georeference. For example, the uncertainty of polygons was measured as the distance from the polygon centroid to the furthest polygon boundary. For certain point and polygon locations where the boundary was not clear, uncertainty was measured as half the distance to the nearest named place. Uncertainty calculations were based on previously published methods ^21,22^.
20. **Diagnostic_Type:** The specific diagnostic test (e.g., polymerase chain reaction [PCR], neutralization test [NT]; hemagglutination inhibition [HI]; enzyme-linked immunosorbent assay [ELISA]) was documented.
21. **Positive_n:** The number of positive MAYV cases that were reported.
22. **Denominator:** The total number of humans, non-human animals, or arthropod pools in the study.
23. **Language:** The language of the article, either English, Portuguese, or Spanish.

### Technical Validation

All georeferencing was completed by one study author and validated by a second author. In the case of a disagreement or discrepancy between the two authors, a third author adjudicated. A location identification was assigned to each unique georeference in the dataset. All final georeferences were plotted in ArcGIS for visual inspection. All data points were checked to ensure they fell on land and within the correct country. In the case of duplicate georeferences, we retained the record with the highest quality score; if quality scores were identical, we retained the more recent record.

### Usage Notes

We identified 144 eligible references for inclusion in our study (see flowchart in Fig 1). The resulting database 266 unique geo-positioned MAYV locations worldwide, including 96 unique points and 170 unique polygons. Therefore, each row in the database represents a unique location where MAYV was detected in humans, non-human animals, or arthropods. Duplicate georeferences from the same host type were removed from the main database (see the Duplicates table in Supplementary material in Data Citation 1) following the approach specified in the methods section. Some duplicate georeferences were included if multiple host types (e.g., human and arthropod) were found with MAYV at the same location. For example, Hoch et al., 1981 ^25^ detected MAYV in both humans and arthropods in the ADM2 unit of Belterra, Brazil. Two separate rows (one row for humans and one for arthropods) are included in the compendium with the same georeference.; therefore, the database includes 280 rows, each representing a unique occurrence of MAYV in a human, non-human animal, or arthropod. Of these 280 rows, 222 (79%) were in humans, 34 (12%) were in non-human animals, and 24 (9%) were in arthropods. MAYV was reported in 15 countries overall, with the majority occurring in Brazil (n = 135). According to our review, MAYV occurrences are limited to the region between latitude 35S and 12N of tropical South America. One article ^26^ reported MAYV occurrence in Zambia; this occurence was not included in our georeferenced database due to the lack of evidence supporting MAYV circulation outside of the Americas and the potential cross-reactivity with antibodies of other alphaviruses in the Semliki Forest serocomplex ^27^.

**Figure 1.**
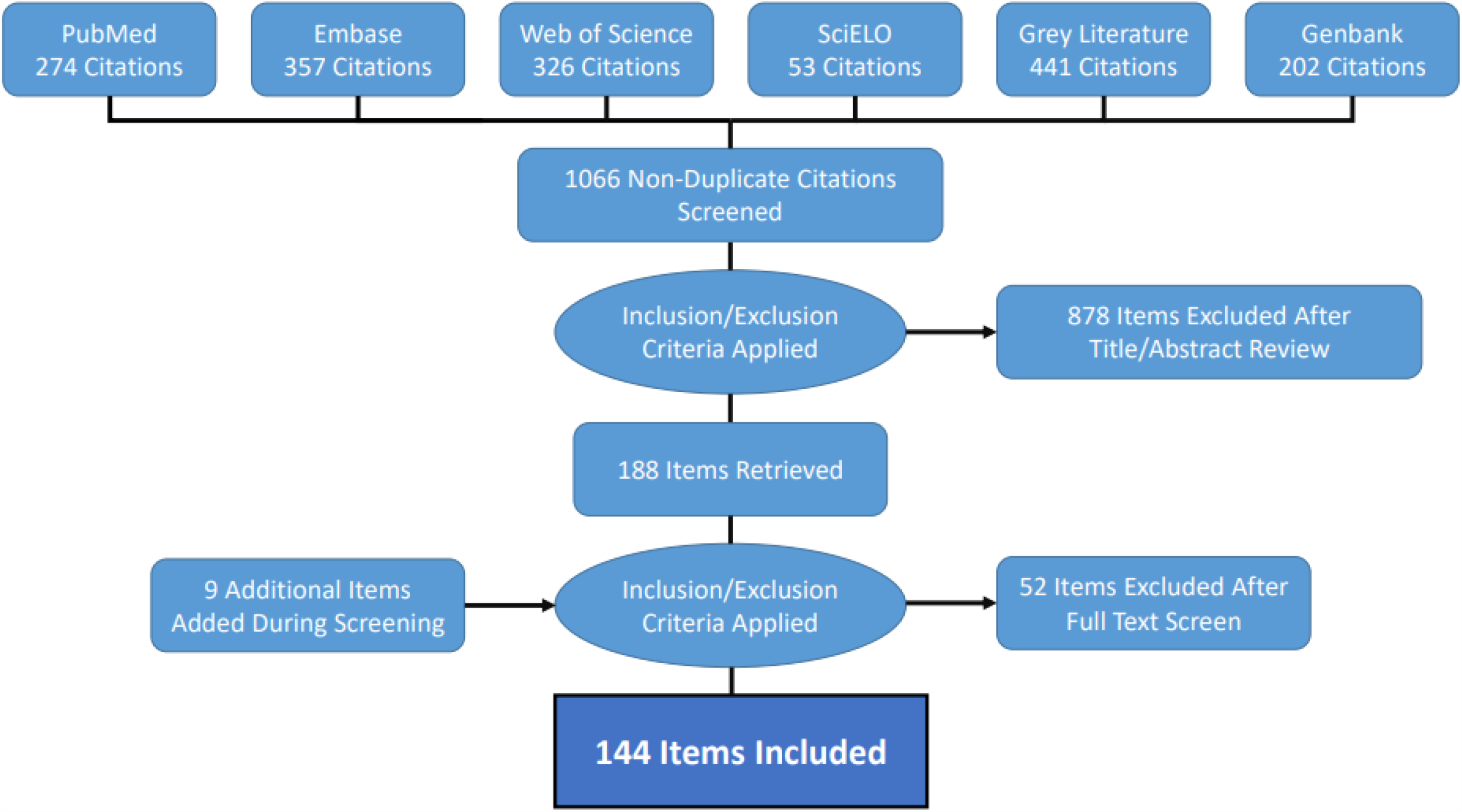
MAYV literature extraction flowchart for human occurrence. The flowchart for non-human animal and arthropod occurrence is provided in the previously published systematic review ^17^.

**Figure 2.**
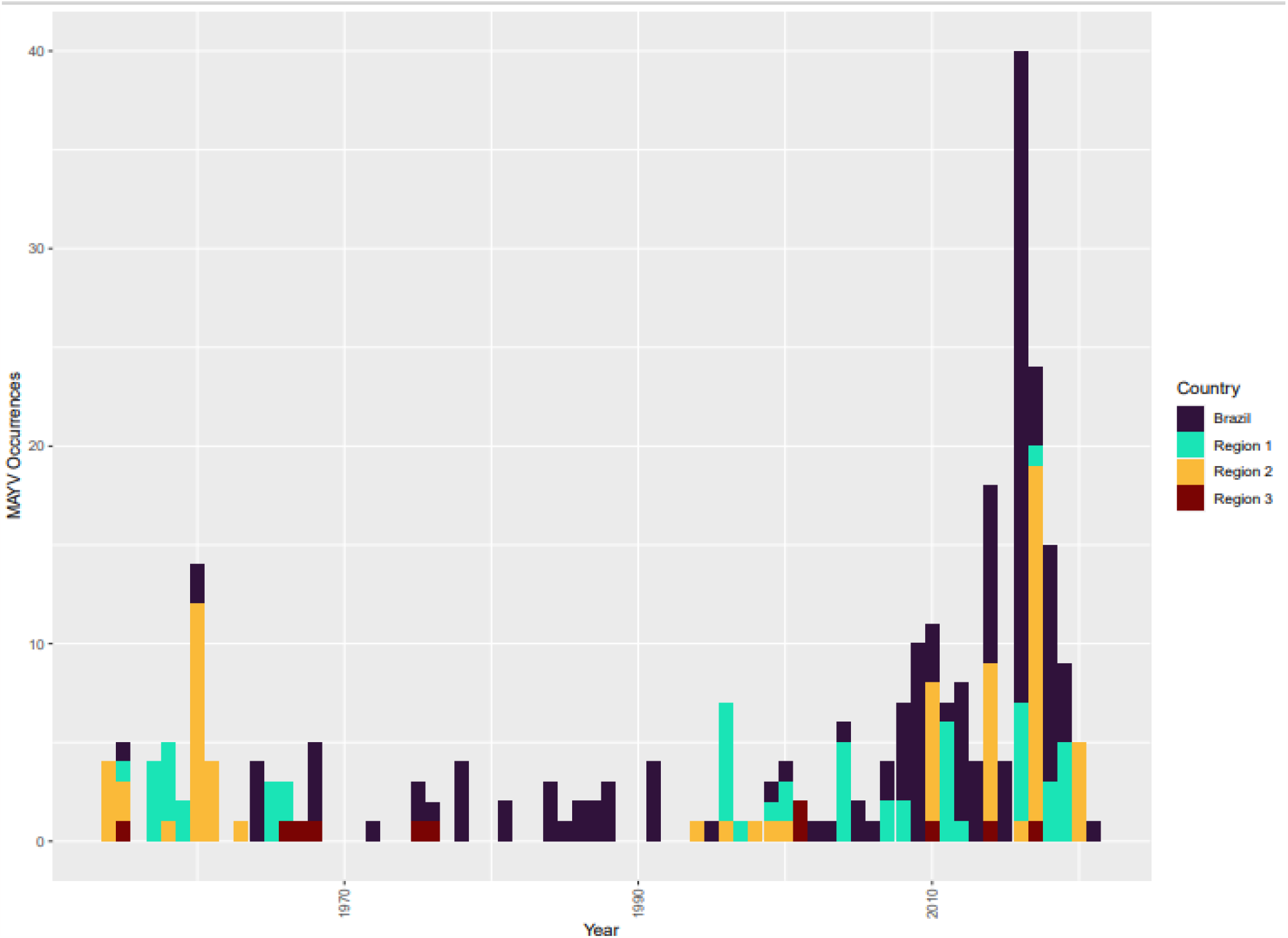
MAYV occurrence (all host types) by year and region. All countries except Brazil were grouped according to geographic region. Region 1 includes Peru, Bolivia, Ecuador, and Colombia. Region 2 includes French Guiana, Guiana, Suriname, Venezuela, and Trinidad & Tobago. Region 3 includes Panama, Costa Rica, Mexico, Haiti and Antigua.

**Figure 3.**
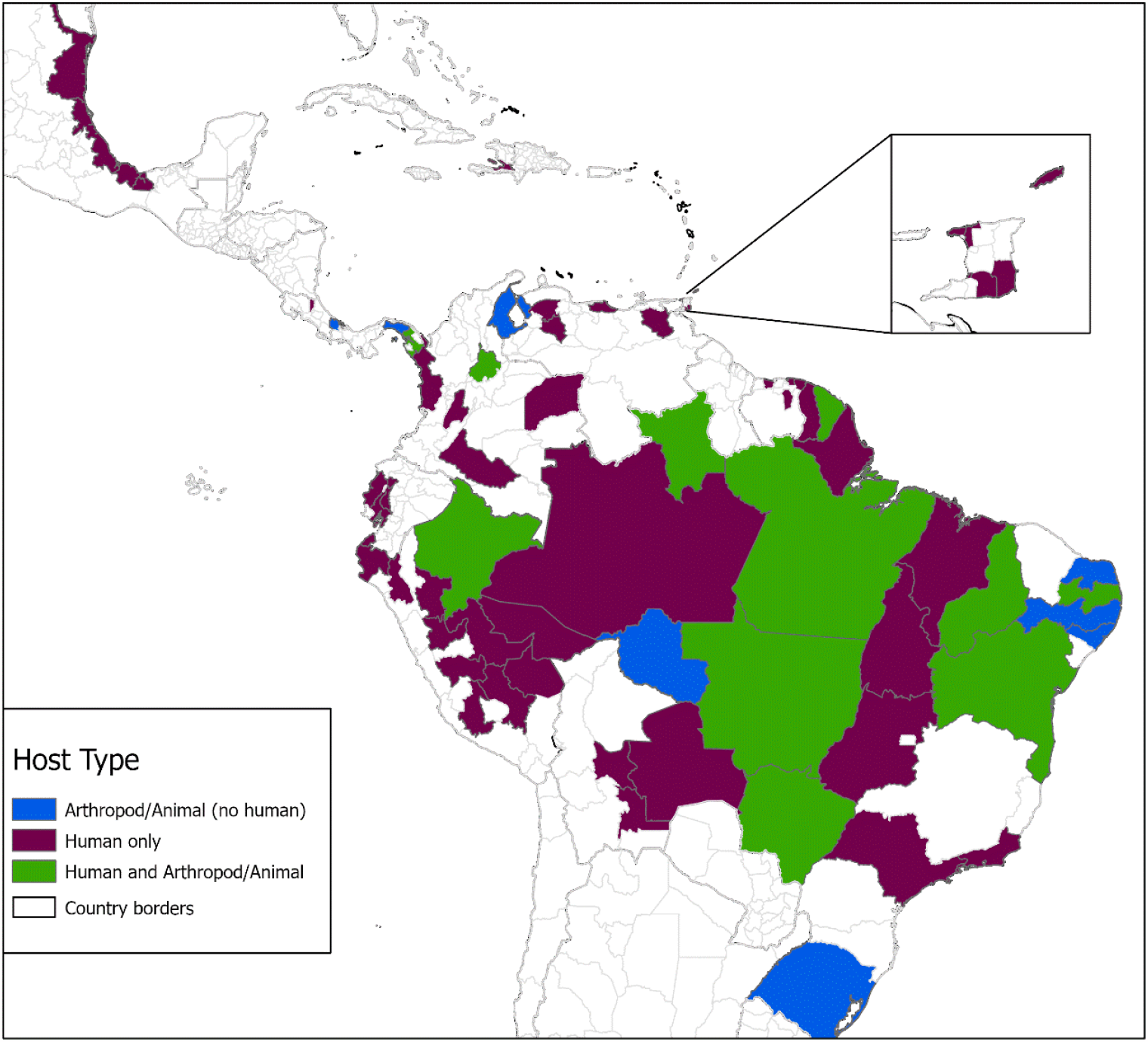
Distribution of MAYV occurrence by first level administrative division. MAYV occurrences are aggregated to the ADM1 level and presented by host type. Host types include human only, reservoir only (non-human animal, arthropod, or both), or human and reservoir (human and non-human animal or arthropod, or all three host types). The inset map shows Trinidad and Tobago.

**Figure 4.**
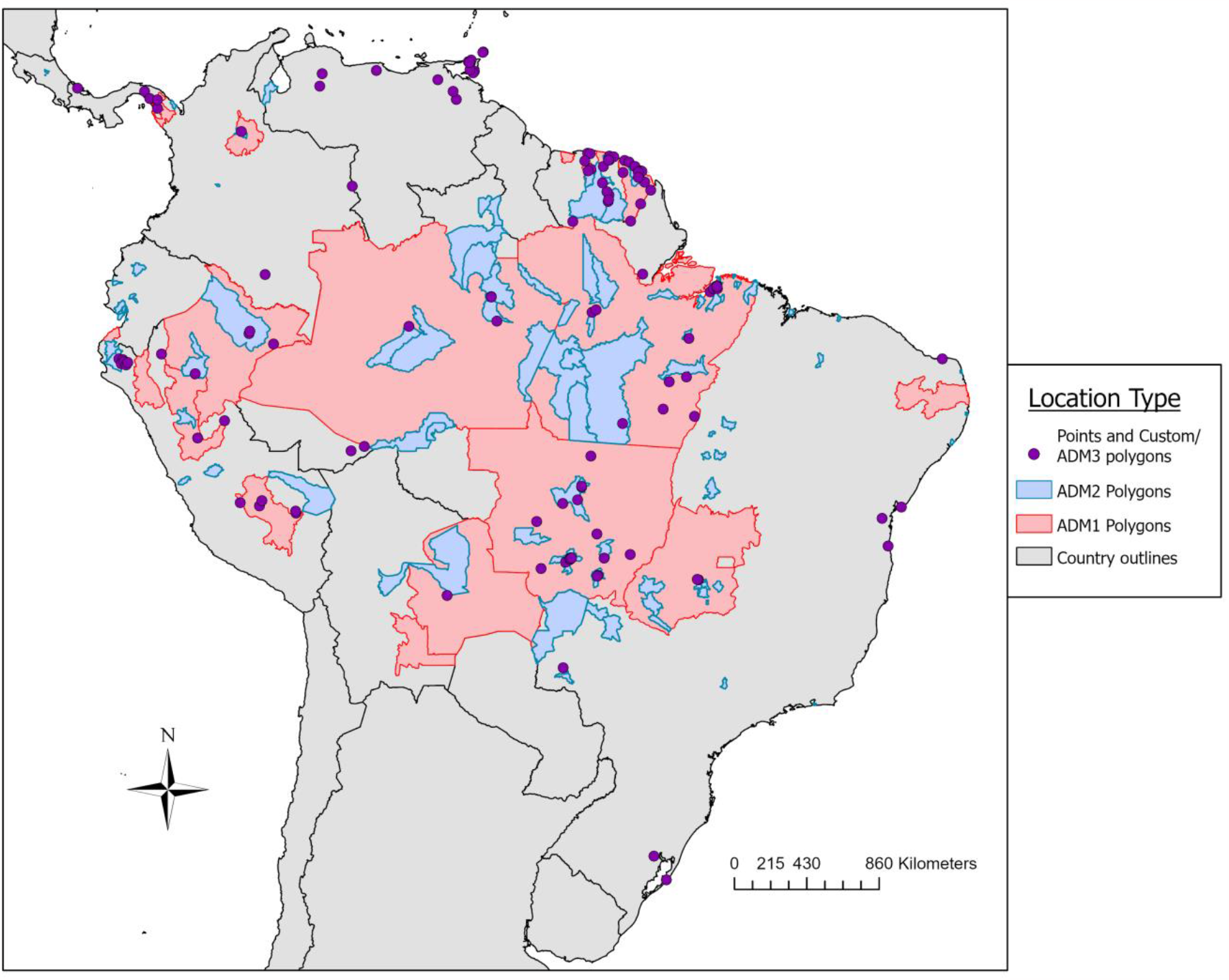
Distribution map of MAYV occurrence by location type. All unique MAYV occurrences are presented according to the precision of the georeference. Red outlines represent first-level administrative units and blue outlines represent second-level administrative units. Both point locations and custom polygons are represented as purple points. Not visible on the map are two ADM2 polygons in Mexico and one ADM2 polygon in Haiti.

Studies were included in our systematic review if they reported testing for MAYV occurrence, even if MAYV was not detected. These negative results are not included in the georeferenced compendium, but they can be found in the Supplementary materials (Data Citation 1). Many machine-learning models that are common in the ecological literature are presence-only or presence-background algorithms that rely on “pseudoabsence” data in lieu of true absences. For this reason, the true absence data presented here are potentially valuable for disease modelling. However, any reports of disease absence must be considered carefully as true absence is difficult to establish and false absence data can result in miscalibration of distribution models ^28^. Ideally, representative country-wide surveys should be used to ascertain “true” absence locations that can be used in subsequent modelling efforts ^29,30^.

As with other published compendiums ^11^, these curated data derived from published sources are expected to complement and augment other survellance data used by public health agencies, thereby increasing our understanding of the distribution of MAYV in the Americas across multiple host types with a high spatial resolution. These data may also assist in identifying under-sampled regions, and may assist in identifying priority regions for surveillance. The georeferences can also serve as the basis for development of epidemiological models or risk maps that characterize the potential suitability for MAYV occurrence, including the risk of spillover into human populations and the potential influence of climate change on MAYV distribution. For example, the 2013 compendium of dengue virus (DENV) occurrence ^12^ was used as the basis for a highly cited modelling study that estimated the global distribution of DENV risk ^31^. Finally, leveraging the methods and data presented here, this open access database can be updated as additional studies are published that report MAYV in the Americas.

There are several important limitations that must be considered when using this dataset. One significant limitation is the impact of sampling bias on the detection and public reporting of MAYV occurrence. Heterogeneity of public health arboviral survellance systems (including variability in surveillance infrastructure and competing public health demands) and MAYV research activity may skew MAYV detection and reporting by geographic region. Therefore, the absence of MAYV occurrence in some settings may not represent true disease absence, but rather ascertainment bias. This important limitation must be addressed in subsequent modeling studies in order to reduce the effects of sampling bias on model accuracy ^32^. Some published studies have proposed an evidence consensus score which quantifies the evidence supporting the presence or absence of a pathogen in a given region ^33^. This score can be calculated using multiple evidence categories (e.g., health organization reporting status or health expenditure) which may provide useful evidence of disease presence or absence in areas, including those with more limited arboviral surveillance.

Another limitation of our study is the lack of geographic precision associated with MAYV occurrence records. Many articles did not provide sufficient geographic detail to georeference MAYV records with a high level of precision. We attempted to capture this uncertainty by assigning polygon locations to these records. When a greater level of geographic detail was provided by study authors, we were able to georeference some records as point locations (i.e., locations of MAYV occurrence with less than 5 km of uncertainty).

Finally, an additional limitation is associated with the variable assay validity used to detect MAYV. Some studies reported MAYV presence based only on positive serological assays such as hemagglutination inhibition (HI) tests while other studies provided stronger evidence of MAYV occurrence based on reference neutralization assays or PCR testing. We estimated the strength of evidence of MAYV occurrence using a custom evidence grade which could be used in other studies. The strength of evidence annotated in these data can be considered in future modeling efforts, with certain low-evidence records potentially excluded from models as part of sensitivity analyses. Moreover, the variability of evidence for MAYV occurrence demonstrated here prompts study design considerations for future MAYV research and public health surveillance.

In addition to the main comma-delimited database, three Supplementary Files are included as part of the file set which can be found online (Data Citation 1). These files include: (i) a document containing the quality score and a citation for each of the references included in our review, (ii) a list of duplicate georeferences that were excluded from the database, and (iii) a shapefile of the custom polygons. The studies that described only negative MAYV results (i.e., those that are not included in the georeferenced compendium) are indicated by an asterisk in the S2 File.

## Data Availability

All data produced are available online in the Dryad repository

https://datadryad.org/stash/share/2EY752eZN2Rfh8LXL6QHXfU6bxHPEjWQcchikr61ImM

## Acknowledgements

The contents, views or opinions expressed in this publication or presentation are those of the authors and do not necessarily reflect official policy or position of Uniformed Services University of the Health Sciences, the Department of Defense (DoD), or Departments of the Army, Navy, or Air Force. Mention of trade names, commercial products, or organizations does not imply endorsement by the U.S. Government.

SP was supported by the National Institute of Allergy and Infectious Diseases, National Institutes of Health, https://www.niaid.nih.gov/, under Inter-Agency Agreement Y1-AI-5072, and the Defense Health Program, U.S. DoD, under award HU0001190002. AP was financially supported by the Armed Forces Health Surveillance Division – Global Emerging Infections Surveillance (AFHSD-GEIS) award P0065_22_WR. The activities undertaken at the Walter Reed Biosystematics Unit were performed in part under a Memorandum of Understanding between the Walter Reed Army Institute of Research (WRAIR) and the Smithsonian Institution, with institutional support provided by both organizations. The funders had no role in study design, data collection and analysis, decision to publish, or preparation of the manuscript.

## Author Contributions

M.C., B.A.H, A.A., and S.P. drafted the data collation protocol. M.C., A.P., B.F., J.D., G.R., N.K.M.M., E.L.L, R.K., I.C.S, K.P, R.C.M, and S.P. assisted with article review, evidence grading, and data extraction. M.C., A.P., N.K.M.M, E.L.L, R.K., and I.C.S contributed to georeferencing. C.L.M. checked all extracted data. M.C. and S.P. wrote the first draft, and all authors contributed to the manuscript.

## Data Citation

To download data click the following link:

https://datadryad.org/stash/share/2EY752eZN2Rfh8LXL6QHXfU6bxHPEjWQcchikr61ImM

## Code Availability

No custom code was used to generate or process the data described in the manuscript.

